# Increasing power in the analysis of responder endpoints in rheumatology: a software tutorial

**DOI:** 10.1101/2020.07.28.20163378

**Authors:** Martina McMenamin, Michael J Grayling, Anna Berglind, James MS Wason

**Affiliations:** MRC Biostatistics Unit, University of Cambridge, Cambridge, United Kingdom; Population Health Sciences Institute, Newcastle University, Newcastle upon Tyne, United Kingdom; Late RIA R&D BioPharmaceuticals, AstraZeneca, Gothenburg, Sweden

## Abstract

**Background:** Composite responder endpoints feature frequently in rheumatology due to the multifaceted nature of many of these conditions. Current analysis methods used to analyse these endpoints discard much of the data used to classify patients as responders; they are therefore highly inefficient and result in low power.

**Methods:** We highlight a novel augmented methodology that uses more of the information available to improve the precision of reported treatment effects. Since these methods are more challenging to implement, we have developed free, user friendly software available in a web-based interface. The software consists of two programs: one that supports the analysis of responder endpoints; the second is used for sample size estimation. We demonstrate the augmented analysis method and its software using the MUSE study, a phase IIb trial in patients with systemic lupus erythematosus.

**Results:** We show the software can be used to analyse the trial with efficiency gains translating to a reduction in required sample size of 63%. Furthermore, we illustrate how the software can be used to choose the sample size needed in a future trial that will use the novel approach as the primary analysis method.

**Conclusion:** We encourage trialists to utilise the software we have developed to implement augmented methodology in future studies to improve efficiency.

## 1. Introduction

Composite endpoints combine a number of individual outcomes in order to assess the effectiveness or efficacy of a treatment. They are typically used in situations where it is difficult to identify a single relevant endpoint to sufficiently capture the change in disease status incited by the treatment, however they may be employed for multiple purposes^1-3^. A subset of these endpoints, known as composite responder endpoints, are commonly used in studies of rheumatic conditions^4-6^. These endpoints allocate patients as either ‘responders’ or ‘non-responders’ based on whether they cross predefined thresholds in the individual continuous outcomes or respond in individual binary outcomes, and are typically treated as a single binary endpoint. A review of core outcome sets across a range of disease areas identified 13 conditions within the domain of rheumatology where an endpoint assuming this structure is recommended to be reported as a primary or secondary endpoint^7^. Table 1 details a typical composite endpoint in each of these 13 conditions along with the response criteria. The endpoints range from single dichotomised measures such as that used in acute gout and vasculitis disorders, to combinations of continuous and discrete outcomes, such as those used in juvenile arthritis, systemic sclerosis and ankylosing spondylitis. As the review considered only outcomes listed within core outcome sets, it is likely a conservative estimate of the number of rheumatology diseases using these measures.

**Table 1:** List of rheumatology conditions where composite responder endpoints containing at least one continuous component are used; *denotes a single dichotomized continuous variable

| Condition | Endpoint | Response definition |
| --- | --- | --- |
| Acute Gout | Proportion of patients who responded* | sUA level of <6.0mg |
| Ankylosing spondylitis | ASAS20 response | 20% improvement and $\geq 10$ units of change (on a 0–100 scale) in each of 3 domains<br>No worsening of a similar amount in the fourth domain<br>(Components are physical function, pain, inflammation and patient's global assessment) |
| Idiopathic arthritis-associated uveitis | Best corrected visual acuity above threshold and no light perception | Best-corrected visual acuity, thresholds $\leq 20/50$, $\leq 20/200$<br>no light perception<br>Estimate contribution of amblyopia, yes/no |
| Juvenile arthritis | Response | Improvement by 30% in at least 3 of:<br>MD global assessment;<br>parent or patient global assessment<br>functional ability;<br>number of joints with active arthritis;<br>number of joints with limited range of motion;<br>Erythrocyte sedimentation rate) |
| Juvenile dermatomyositis | Responder index | $\geq 4$ point reduction from baseline in safety of estrogen in lupus national assessment (SELENA) systemic lupus erythematosus disease activity index (SLEDAI) score |
|  |  | • No worsening (increase of <0.30 points from baseline) in physician's global assessment (PGA)<br>• No new British Isles Lupus Assessment Group of SLE clinics (BILAG) A organ domain score or 2 new BILAG B organ domain scores compared with baseline |
| Prevention of fracture in high risk populations | Response | • Bone mineral density increase<br>• Occurrence of new vertebral fractures |
| Proliferative and membranous lupus renal disease | Urinary protein levels within normal range* | • Between 6 and 8.3 grams per deciliter (g/dL) |
| Rheumatoid arthritis | ACR20 response | • $\geq 20\%$ improvement in ACR score<br>• Can be combined with additional requirements e.g. no additional medication |
| Sarcopenia prevention | Occurrence of sarcopenia | Heterogeneity in precise definition, but severe sarcopenia defined by all of the following:<br>• Low muscle strength (assessed with chair stand test or grip strength)<br>• Low muscle quantity/quality<br>• Low physical performance as assessed with gait speed test or short physical performance battery |
| Sjogren's syndrome | Response | • >30% reduction in analog scales evaluating dryness, pain and fatigue |
| Systemic lupus erythematosus | SRI responder index | • SLEDAI change e.g. $\leq -4$<br>• PGA change e.g. $<0.3$<br>• No Grade A or more than one Grade B in BILAG |
| Systemic Sclerosis | SCP in normal range, no renal crisis | E.g.<br>• $< 3.0\text{mg/dl}$ not drug related<br>• No renal crisis |
| Vasculitis disorders | Response/partial improvement* | • 50% improvement in disease activity score. |

Employing composite endpoints as the primary outcome measure in a study has many advantages. Proponents of composite endpoints believe that they are appropriate as they estimate the net clinical benefit of an intervention by accounting for the multiple factors of interest in a given disease^8-10^. This is especially important in complex, multisystem, chronic diseases typical in rheumatology, to ensure that while a patient may have improved overall on one scale, that a flare in another organ domain is not introduced on another scale. Furthermore, in the case of diseases with large variation in symptoms, employing a composite endpoint will avoid an arbitrary choice of a single outcome^11-12^. However, many problems with the application of composite endpoints have been raised in the literature. In practice, composites may be inconsistently defined and provide opportunities for post-hoc changes^13^. Composite endpoints may also be driven by less important or subjective components, meaning that a promising treatment effect may not translate to benefit for patients. Moreover, they have the tendency to become very complicated and therefore difficult for physicians and patients to understand.

Additional criticisms arise from the analysis of these endpoints. The endpoints are typically treated as binary measures based on whether or not the patient responded, meaning the analysis is straight-forward to implement. However, for composites containing continuous outcome measures, this is at the expense of losing large amounts of information contained in those components^14^. In the context of phase II oncology responder endpoints, Wason and Seaman^15^ proposed a novel technique to address these issues, utilising a more complex model to retain information on how close patients were to the response thresholds in the continuous measures. This has since been developed to include different types of endpoints^16-18^ and for application in rare diseases^19^. It has also been successfully applied retrospectively in trials, including in rheumatoid arthritis^20^ where the efficiency gains translated to a reduction in required sample size of at least 30% and systemic lupus erythematosus (SLE), where the resulting required sample size was reduced by 60%.

One limitation of these methods is that they are more difficult to implement. Therefore, in this paper we demonstrate the use of free, user friendly online software for conducting analyses of composite responder endpoints using the augmented approach. We illustrate this using the MUSE trial^21^, which assessed the efficacy of anifrolumab in patients with SLE. Furthermore, we show how a second software tool may be used to establish the required sample size for a future study in SLE.

The paper proceeds as follows: in Section 2 we give a brief description of the methods, Section 3 summarises the MUSE trial data, Section 4 demonstrates the capabilities of the software and Section 5 discusses the implications for practice.

## 2. Analysis Methods

### 2.1 Standard binary approach

We refer to the analysis method routinely applied to composite responder endpoints as the binary approach. This consists of collapsing the outcome information to form a binary response variable based on whether or not the patients meet the overall response criteria. This response variable is analysed using an appropriate binary analysis method, such as logistic regression. The treatment effect can then be reported in terms of odds ratios, risk ratios or risk differences along with confidence intervals and p values.

### 2.2 Augmented approach

The augmented approach involves using a more sophisticated model that jointly models data from each of the components. The information contained in the continuous components is retained and used to weight patients differently in the analysis, based on how close their continuous readings were to the response threshold. The probability of response in each arm is subsequently obtained which can then be used to form treatment effect estimates in terms of odds ratios, risk ratios or risk differences, as in the standard binary case. The increased efficiency compared to the binary approach is due to making inference on the probability of response without discarding any of the continuous data. In datasets where many patients’ continuous readings are close to the dichotomisation threshold, this may have a substantial impact on the precision of the estimate and hence on the conclusions reached.

## 3. MUSE trial summary

To illustrate how the analysis can be conducted using the software, we focus on the MUSE trial^21^. The trial was a phase IIb, randomised, double-blind, placebo-controlled study investigating the efficacy and safety of anifrolumab in adults with moderate to severe SLE. Patients (n=305) were randomised (1:1:1) to receive anifrolumab (300mg or 1000mg) or placebo, in addition to standard therapy every 4 weeks for 48 weeks. The primary end point in the study was the percentage of patients achieving an SLE Responder Index (SRI) response at week 24, with sustained reduction of oral corticosteroids (<10mg/day and less than or equal to the dose at week 1 from week 12 through 24), which is typically referred to as ‘SRI+OCS’. As detailed in Table 1, SRI is comprised of a continuous Physician’s Global Assessment (PGA) measure, a continuous SLE Disease Activity Index (SLEDAI) measure and an ordinal British Isles Lupus Assessment Group (BILAG) measure^22^.

Table 2 shows the decomposition of responders and non-responders in each of the components by treatment arm. In both the treatment and the control arm, almost all patients are responders in both the PGA and BILAG measures. This indicates that these components do not enrich the composite endpoint in this study and so it is the SLEDAI and taper measures that are responsible for driving response rates. Previous work has shown that we may expect smaller efficiency gains than if three or four components determined response^23^. For the purposes of this analysis we combine the ordinal and binary components to form a single indicator as modelling the ordinal component directly vastly increases computing time for only a small increase in efficiency^16^.

**Table 2:**
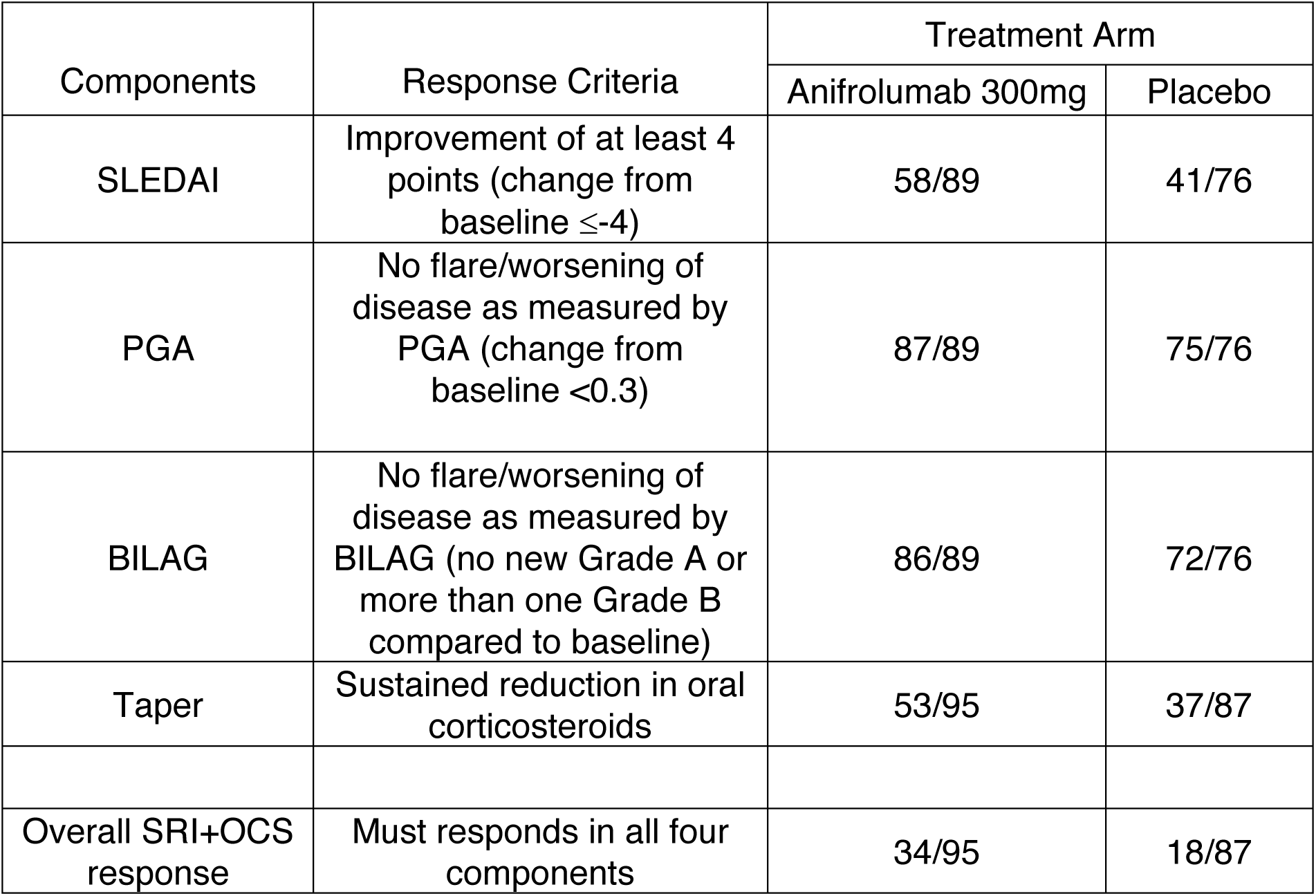
Observed response rates in each of the SRI+OCS components in the anifrolumab 300mg arm and placebo arm of the MUSE trial. SLE index is comprised of a continuous SLEDAI outcome, continuous PGA outcome, ordinal BILAG outcome and binary

## 4. Software Tutorial

### 4.1 Analysis

The software to implement the analysis is a Shiny application, a Graphical User Interface (GUI) for programming language ‘R’ which can be accessed at https://martinamcm.shinyapps.io/augbin/. Underlying code and documentation is available as indicated on the homepage.

The user begins by selecting the analysis tab and uploading the csv file using the ‘Upload Files’ panel. A table displaying the uploaded data will be shown on the right hand side as shown in Figure 1. Note that the data displayed in Figure 1 is not the real data, due to patient confidentiality but that the real trial data is used in what follows. In order to conduct the analysis, the user must organise the columns in the dataset prior to uploading, so that patient ID comes first, followed by treatment arm, the continuous outcomes, the binary outcome and the baseline measures for the continuous variables. Failure to upload a dataset in this format will cause issues at the analysis.

**Figure 1:**
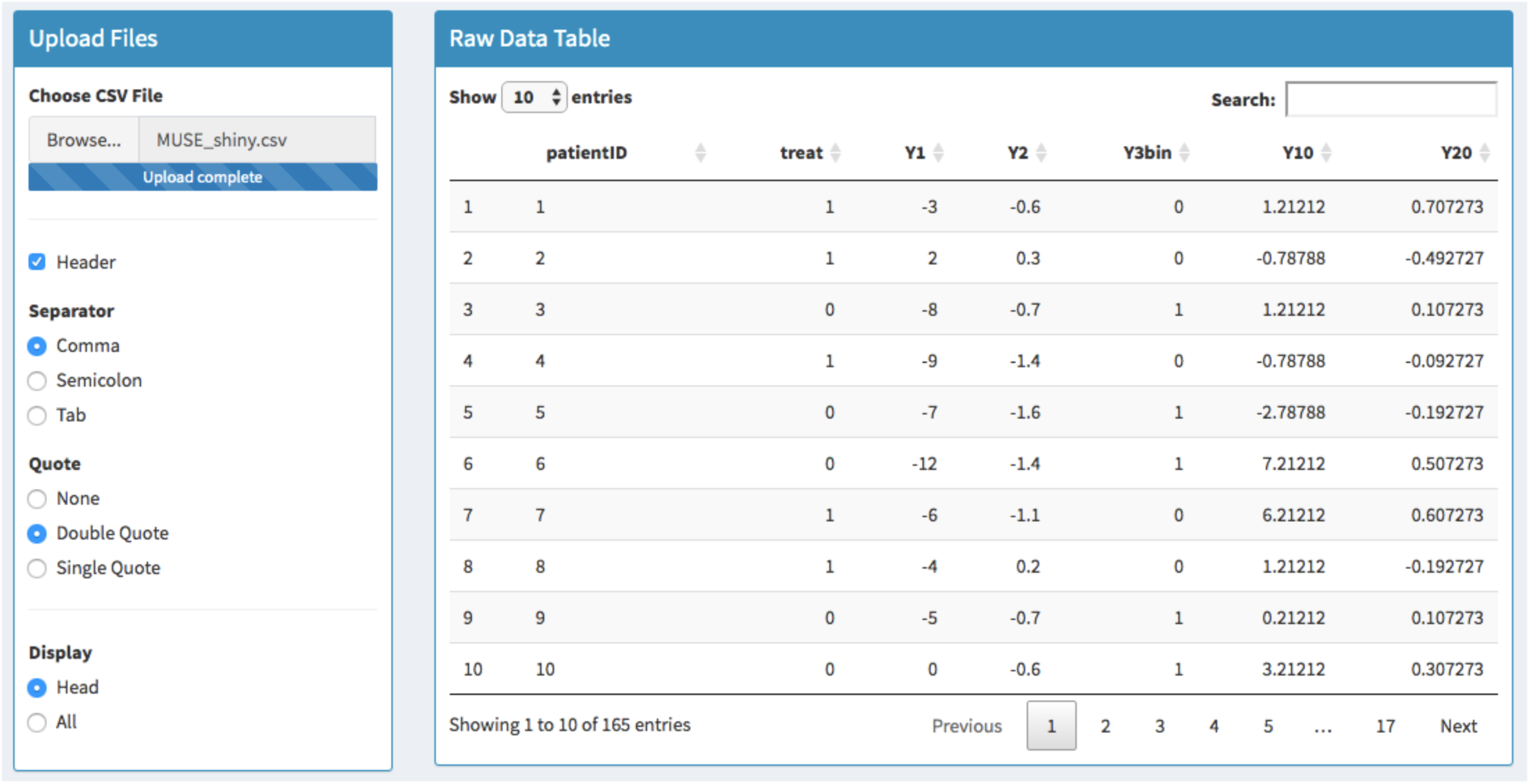
MUSE trial data is uploaded in the left hand panel where the user can indicate preferences such as whether the file includes column headers and whether to display some or all of the data. The raw data is viewed in the right hand panel where users may also search for particular subjects.

The raw data can be visualised using boxplots, histograms, density plots or bar graphs in the ‘Raw Data Plots’ panel. The user must then select the structure of the composite endpoint, where this can be one or two continuous components and zero or one binary components. The SLE endpoint has two continuous and one binary components and so the relevant underlying model can be viewed by selecting ‘Generate model’. Both of these steps are demonstrated in the supplementary material.

The analysis is initiated in the ‘Analysis’ panel by selecting the response threshold for the continuous outcomes, where in this case the SLEDAI threshold is -4 and the PGA threshold is 0.3. Figure 2 shows the probability of response in each arm of the MUSE trial using both the novel latent variable approach and the standard binary approach. The log-odds ratio, log-risk ratio and risk difference treatment effects are shown for each method along with the 95% confidence intervals. The augmented approach reports each of the treatment effects more precisely. To gain a similar level of precision whilst using the binary analysis method, one would need to increase the sample size by 270%. The ‘Goodness-of-fit’ panel below indicates how well the more sophisticated latent variable model fits the data, where a ‘good fit’ is assumed if the residuals follow the chi-squared distribution shown by the red line. The goodness-of-fit plots for the MUSE dataset are shown in the supplementary material.

**Figure 2:**
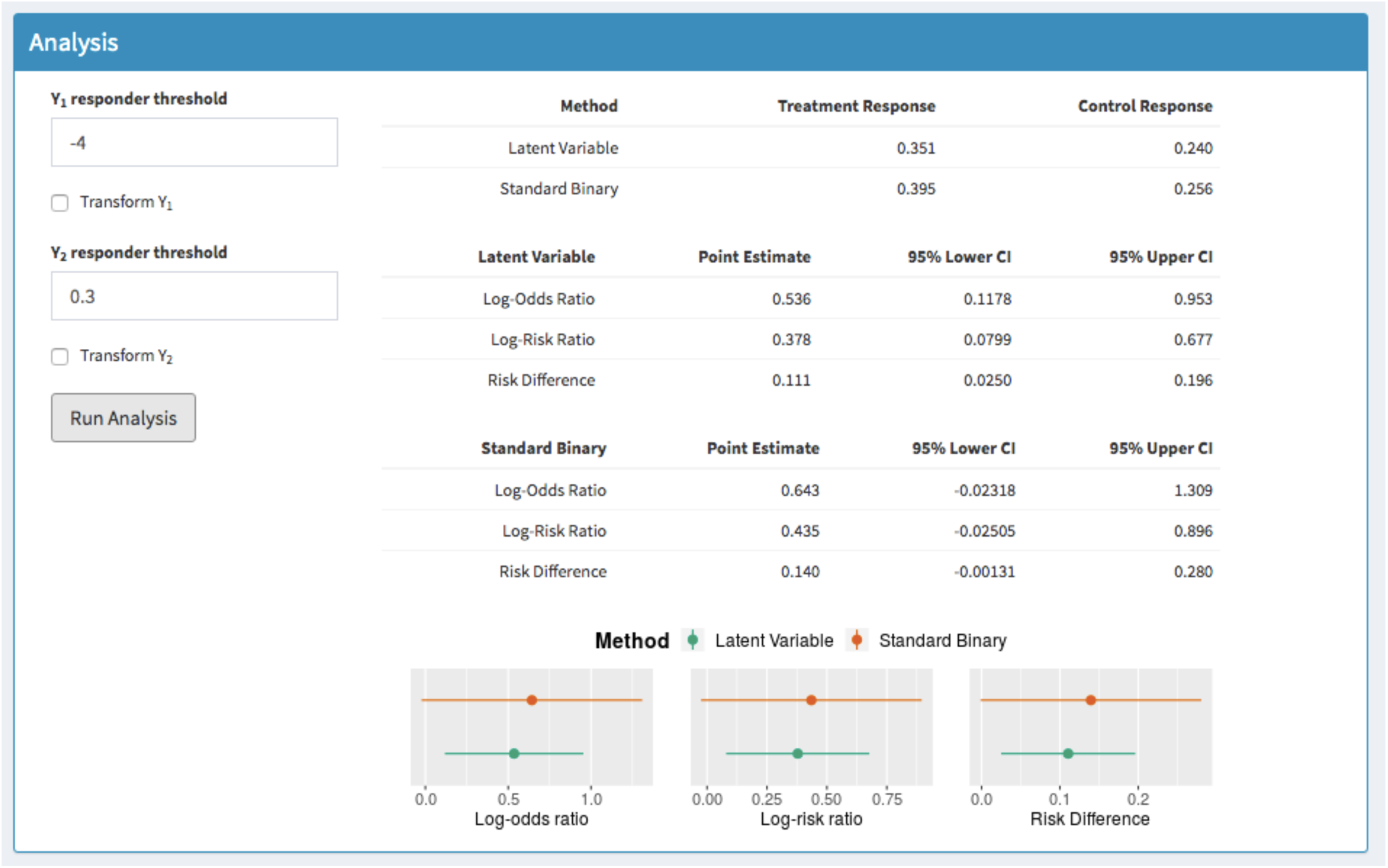
Analysis of the SRI+OCS endpoint in the phase II MUSE trial where the tables show the probability of response in each method, the treatment effects and 95% CIs for the latent variable method and the treatment effects and 95% CIs for the standard binary. The corresponding plots are shown below.

### 4.2 Sample Size Determination

A critical aspect of planning a future study using the augmented approach is how to determine the sample size, in order to avail of the efficiency gains. The ‘MultSampSize’ shiny application allows users to determine sample sizes required through using preliminary data to inform the estimates. This may be in the form of pilot trial data, trial data from earlier phase studies or another source. The sample size estimation app can be accessed at https://martinamcm.shinyapps.io/multsampsize/, where detailed documentation is also included. In order to demonstrate its capabilities, we can assume that we wish to design a future trial in SLE which will use the augmented approach as the primary analysis method. This will be directly informed by estimates from the MUSE study.

The user should select the ‘Sample Size’ tab and choose the ‘Composite’ option to proceed. Note that the app also accommodates co-primary and multiple primary endpoints, which also feature in rheumatology^23^. The user must select the number of continuous and binary components and the corresponding response thresholds, as before. Selecting ‘Get Model’ displays the relevant model assumed along with the power function used to determine the sample size.

Figure 3 shows this for the SLE composite where the assumed primary endpoint is SRI(4) combined with whether the patients can sustain a reduction in oral corticosteroids. The pilot data can be uploaded using the ‘Parameter Estimates’ panel, where the columns must be ordered as before. Further guidance is available at https://github.com/martinamcm/MultSampSize. Clicking ‘Obtain Estimates’ runs the analysis and provides estimates for the probability of response in each arm, the risk difference and its variance. These values are also provided assuming the standard binary method was used, in the ‘Binary’ column as shown in Figure 4. The ‘Sample Size Estimation’ panel displays the power curve and highlights the number of patients needed per arm to attain a desired power and alpha level, which can be set by the user. Assuming a one-sided test with alpha level 0.05, a target power of 80% and using the observed risk difference from the latent variable approach in the MUSE trial, dictates a required sample size of 45 individuals per arm for a future SLE study, compared with 73 patients per arm that would be required using the standard method. Using the observed treatment effect estimate in the standard approach instead would require 28 versus 45 individuals to be enrolled on each arm. The power curve for both the augmented and binary approaches is shown in Figure 4.

**Figure 3:**
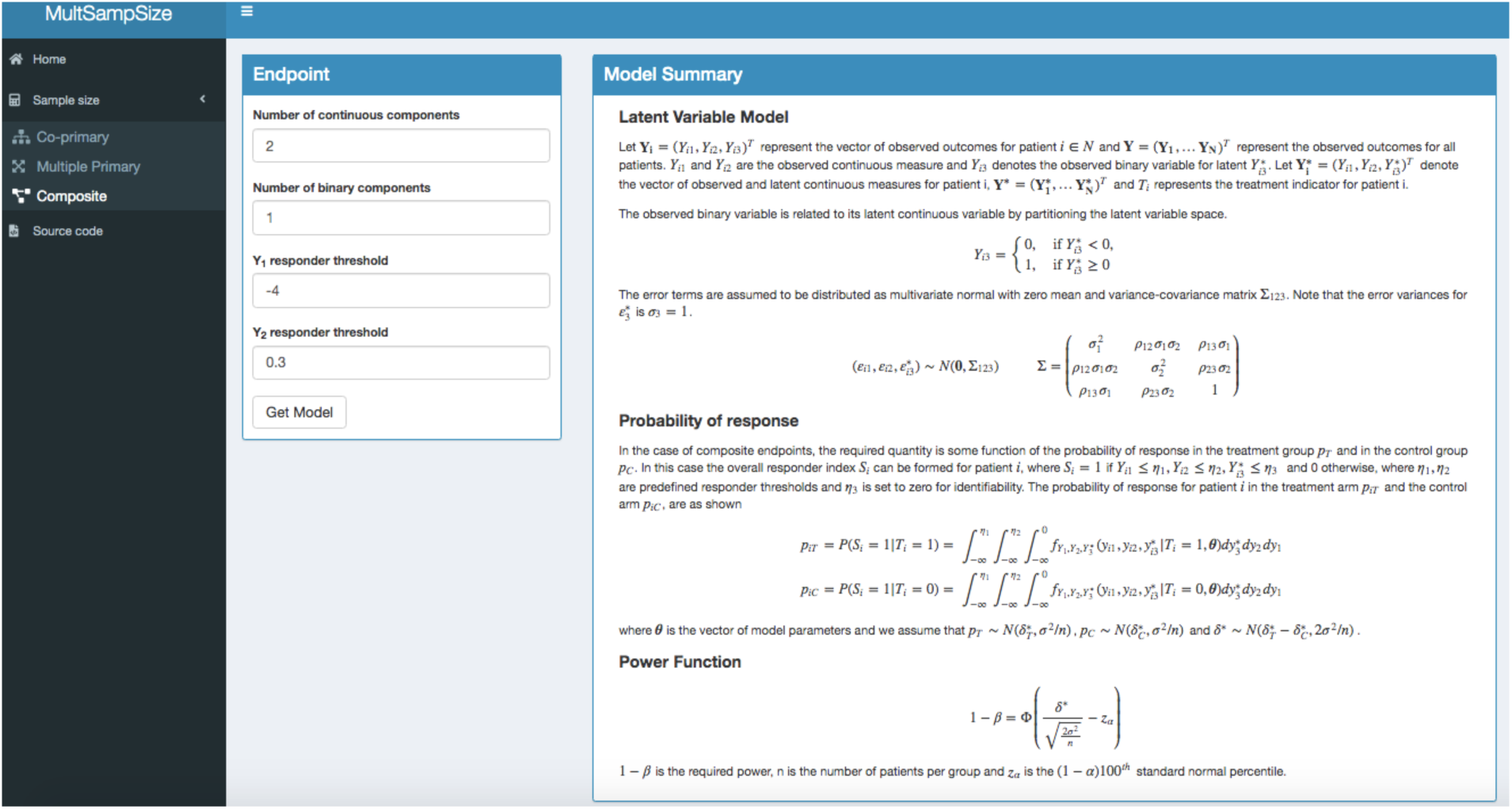
Inputting dichotomisation threshold in ‘MultSampSize’ app

## 5. Discussion

In this paper we highlight novel methods to address inefficiencies in the analysis of composite responder endpoints commonly used in rheumatology, which use more sophisticated models to retain the information provided by continuous outcomes. As this approach is more difficult to implement we developed user friendly, free to use software. This software conducts the analysis using both methods and offers sample size determination for future studies using the augmented technique as the primary analysis method. We demonstrated the functionality of the apps using the MUSE trial dataset, a phase II study in patients with SLE using a composite comprised of two continuous and one binary outcomes.

The analysis took approximately 5 minutes to complete, where the gains in efficiency equated to a 63% reduction in required sample size. This means that 63% fewer patients could be recruited by using the augmented analysis method without requiring any additional data to be collected. Using the MUSE trial to inform a future study in SLE indicated that the novel approach would require 45 per arm versus 73 required for the standard approach.

As the structure of the composite endpoints vary substantially and may be quite complex, the efficiency gains offered by this technique also depends on many factors. In particular, the number of continuous and binary components, response probabilities in each arm, the responder thresholds and correlation between components. Both Shiny applications therefore report the results for the standard and novel approaches. In the case of sample size estimation, the investigator has the option to recruit the number of patients dictated using the binary approach and benefit from the additional power instead.

The methods underpinning the apps allow for any number of continuous, ordinal and binary components to be included in the composite endpoint however the app currently only implements this for up to two continuous components. Each additional continuous endpoint may add a substantial amount of efficiency and so future work will involve updating the software to allow for more complex endpoints such as those used in juvenile arthritis. In its current form the software may still be used for such endpoints however the additional components will have to be combined as a binary indicator. In this case, the most informative continuous outcomes should be retained.

## Data Availability

The authors do not own the trial data used in the manuscript to illustrate the software, so we are unable to make it available.

